# Intraoperative laryngeal muscle and heart rate responses to implanted vagus nerve stimulation

**DOI:** 10.1101/2025.10.03.25337063

**Authors:** Kathryn M. Turk, Kevin J. Mohsenian, Jennifer J. Peters, Derek G. Southwell, Warren M. Grill, Nicole A. Pelot

## Abstract

1.

**Objectives:** Implanted cervical vagus nerve stimulation (VNS) is used to treat refractory epilepsy, depression, stroke sequelae and rheumatoid arthritis. The therapeutic efficacy of VNS is limited by stimulation-induced side effects, including hoarseness, coughing, and voice alteration. We quantified VNS-evoked laryngeal muscle activation (EMG; indicating side effects) and changes in heart rate (HR; proxy for activation of therapeutic fibers) in participants undergoing VNS implant surgery.

**Methods:** We recruited adult participants (7F/3M) with treatment-resistant epilepsy who were receiving a new VNS implant (“acute”) or replacement of an implanted VNS pulse generator (“chronic”). During these procedures, we delivered VNS across pulsewidths (50, 250, and 1000 µs/phase) and stimulation amplitudes (0.05-28 mA) while recording laryngeal EMG and HR.

**Results:** The median stimulation amplitudes to evoke 50% of maximal laryngeal EMG response were 1.32, 0.49, and 0.34 mA for pulsewidths of 50, 250, and 1000 µs/phase, respectively; thresholds were comparable between EMG electrodes placed endotracheally and subcutaneously. The median stimulation amplitudes to cause a 10% decrease in HR were 13.39 and 3.53 mA at 50 and 250 µs/phase, respectively—i.e., ~6 to 63x higher than the 50% EMG thresholds. We did not observe a difference in EMG or HR responses between sexes, acute/chronic, or stimulation polarities. For each subject in the chronic implant group, clinician-selected stimulation amplitudes were higher than the 50% EMG thresholds and lower than the 10% HR thresholds.

**Conclusions:** Thresholds to evoke bradycardia were ~2x higher than clinician-selected stimulation amplitudes. This indicates that the target fibers of VNS for epilepsy may be larger diameter than those projecting to the heart. Alternatively, the therapeutic effect in refractory epilepsy may be evoked with less fiber activation than is required to produce bradycardia.

## 2. Introduction

Implanted cervical vagus nerve stimulation (VNS) is used clinically to treat refractory epilepsy (Englot et al., 2017), depression (Aaronson et al., 2017), stroke sequelae (Dawson et al., 2021) and rheumatoid arthritis (Koopman et al., 2018; Peterson et al., 2024), and VNS is under investigation to treat several other conditions, including heart failure (Gold et al., 2016), and Crohn’s disease (Bonaz et al., 2017). However, the therapeutic efficacy of VNS is limited by side effects, including hoarseness, coughing, dyspnea (shortness of breath), and voice alteration (Milby et al., 2009).

Most VNS side effects are caused by activation of large diameter motor fibers in the vagus nerve that innervate laryngeal muscles. Prior studies recorded laryngeal muscle responses in response to VNS (Ardesch et al., 2010, 2007; Banzett et al., 1999; Binks et al., 2001; Bouckaert et al., 2022; Chiba et al., 2019; Lundy et al., 1993; Saibene et al., 2017; Shaffer et al., 2005), and thus, laryngeal muscle responses can serve as a biomarker of vagal activation (Chang et al., 2020; McAllen et al., 2018; Vespa et al., 2019). The fibers that innervate laryngeal muscles are large diameter A-alpha motor fibers, whereas therapeutic fibers are suggested to be smaller diameter myelinated afferent and/or efferent fibers (Ardell et al., 2017; Yoo et al., 2013) which have higher thresholds for activation by VNS.

The heart is innervated by small, myelinated parasympathetic efferent fibers of the vagus nerve, and their activity induces bradycardia (i.e., slowing of the heart rate (HR)). Prior studies measured HR during VNS (Ardesch et al., 2007; Banzett et al., 1999; Binks et al., 2001; Frei and Osorio, 2001; Premchand et al., 2014), but no data exist reporting thresholds for evoking bradycardia with VNS in humans (Musselman et al., 2023b). Indeed, three pivotal clinical trials of VNS for heart failure failed to reach their efficacy end point, and stimulation did not evoke bradycardia—suggesting that the targeted therapeutic small, myelinated fibers were not activated—whereas bradycardia was evoked by VNS in preclinical experiments (Musselman et al., 2019). Bradycardia is not required for efficacy in treating heart failure, but it is a measurable effect of activation of small, myelinated parasympathetic fibers in the vagus nerve.

Fiber activation thresholds in response to peripheral nerve stimulation are influenced by multiple factors, including fiber diameter, electrode-fiber distance, and fascicle diameter (Davis et al., 2023). However, the functional organization of fibers in the human vagus nerve is unknown, and thus, the relationship between VNS activation thresholds of large versus small, myelinated fibers is also unknown. Further, the fascicular anatomy of the human vagus nerve is highly variable between individuals (Pelot et al., 2020). Quantification of the differential activation thresholds for laryngeal muscles (side effects) and bradycardia (proxy for on-target fibers) will provide important insights into the direct neural responses to VNS.

In this study, we measured VNS thresholds intraoperatively. We recruited adult participants with epilepsy who underwent surgery either for an initial implant of a VNS device or for replacement of the implanted pulse generator (IPG) due to battery depletion. During surgery, we evaluated the effects of VNS parameters (pulse duration, pulse amplitude) on laryngeal muscle activation and changes in HR as proxies for activation of large and small, myelinated fibers, respectively.

## 3. Methods

We recruited n = 14 adult participants with treatment-resistant epilepsy who were either receiving a new VNS implant (“acute”) or a replacement IPG (“chronic”) as part of their standard of care. During their surgery, we conducted 30 minutes of intraoperative research. We connected research stimulation equipment to their VNS electrode lead to stimulate with different pulsewidths (50, 250, 1000 µs) and amplitudes (0.05-28 mA). We also evaluated the effects of stimulation polarity, and in chronic participants, we recorded responses to clinically programmed parameters. We recorded laryngeal muscle responses (EMG) via endotracheal and needle electrodes, and we recorded cardiac effects via electrocardiogram (EKG).

We successfully collected data from ten participants (4 chronic female, 3 acute female, 2 chronic male, 1 acute male), including one participant from whom only EMG data were recorded due to time constraints (1 acute female, P5). Data were not collected from four participants due to technical issues.

### 3.1 Participants and Survey

All participants were adults (18-65 years old, inclusion criterion), neurologically stable apart from epilepsy and provided informed consent. Individuals with another implanted device or a history of heart conditions that the surgeon determined to be a risk were not recruited. All protocols were approved by the local Institutional Review Board, and the trial was registered with clinicaltrials.gov.

Each participant completed a pre-surgical survey (Supplement I) to interrogate their current seizure frequency and quality of life. For chronic participants, the survey additionally asked about their experience with VNS, including change in seizure frequency, change in seizure quality, and side effects. For all participants, we recorded the model of the implanted IPG and the impedance of the VNS electrode at the end of the case (Supplement II). For chronic participants, the model of the removed IPG and time from initial implant were also recorded.

### 3.2 Instrumentation

Prior to surgery, participants were intubated with a Medtronic NIM-EMG endotracheal tube (product #8229707 or #8229706, Medtronic; Minneapolis, MN), which has two pairs of electrode contacts to record laryngeal EMG (Figure 1). A short-acting paralytic (succinylcholine) was delivered just prior to intubation; its rapid washout enabled recording of EMG during research. Anesthesia was induced and maintained with propofol and fentanyl. All participants were mechanically ventilated. Before the first incision, five needle electrodes (13 mm, product #12.5RSLD, Rhythmlink; Columbia, SC) were placed subcutaneously (Figure 1): one needle was placed on either side of the laryngeal prominence (~2-3 cm from midline) to record laryngeal EMG, and three needles were placed in the left and/or right shoulder to serve as references for the EMG amplifiers.

**Figure 1.**
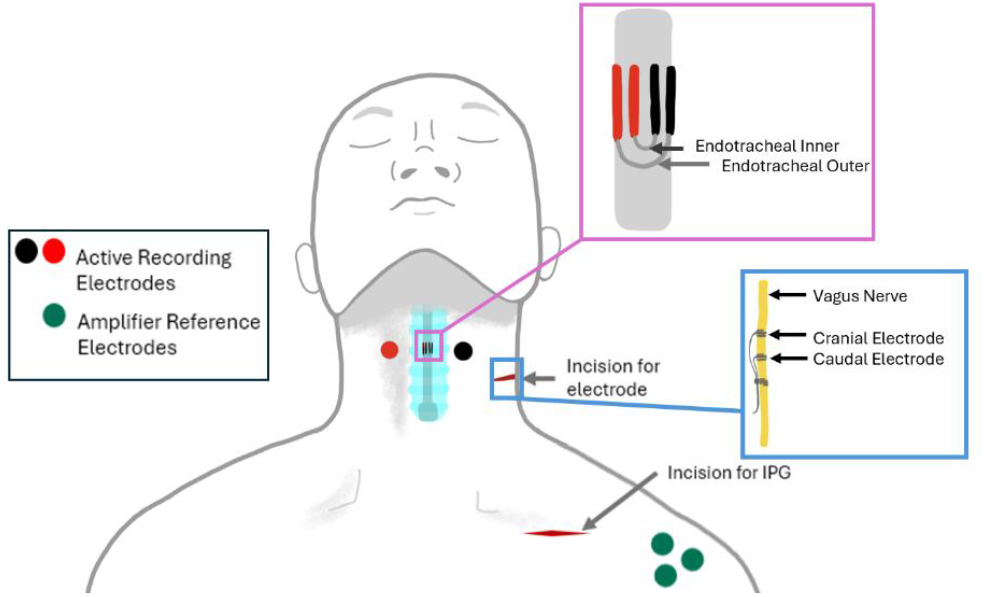
Laryngeal EMG recording electrodes, showing electrode locations for needles (circles) and the endotracheal tube (inside trachea shown in blue). The purple inset shows the connection of inner and outer pairs of channels for the endotracheal recording. Red electrodes denote the non-inverting input to the differential amplifier and black electrodes denote the inverting input. Green circles denote the three reference electrodes for the active recordings (needle and endotracheal tube electrodes). The blue inset shows the VNS electrode. IPG: implanted pulse generator.

A LivaNova VNS device was used in all cases, with a 2 mm-diameter cuff electrode; all cases received the Sentiva 1000 IPG, while some chronic cases previously had a different IPG model (Supplement II). In acute cases, the cuff electrode was first placed on the left cervical vagus nerve, then the leads were tunneled to the chest. In chronic cases, the implanted IPG was first removed from the left chest. For both acute and chronic cases, the VNS electrode lead was then connected to a sterile extension cable (ATAR D-RL, OSCOR Europe GmbH, Düsseldorf, Germany), and surgery was paused for research testing (30 min). The extension cable was connected such that the cathodic-leading stimulation pulse was delivered to the cranial contact, consistent with clinical VNS for epilepsy and depression.

Figure 2 diagrams the instrumentation. Stimulation parameters were defined in MATLAB (R2022b, MathWorks; Natick, MA) and delivered via a National Instruments Data Acquisition System (NI-DAQ, USB 6216; Austin, TX) which provided a voltage-controlled signal to a DS5 Isolated Bipolar Current Stimulator (Digitimer; Hertfordshire, UK). The current-controlled stimulation signal was then filtered to remove any contaminating direct current (dc) before delivery to the VNS electrode lead (Supplement III: Figure 10). The voltage-controlled signal from the NI-DAQ and the DS5 “monitoring” output were both recorded by the NI-DAQ.

**Figure 2.**
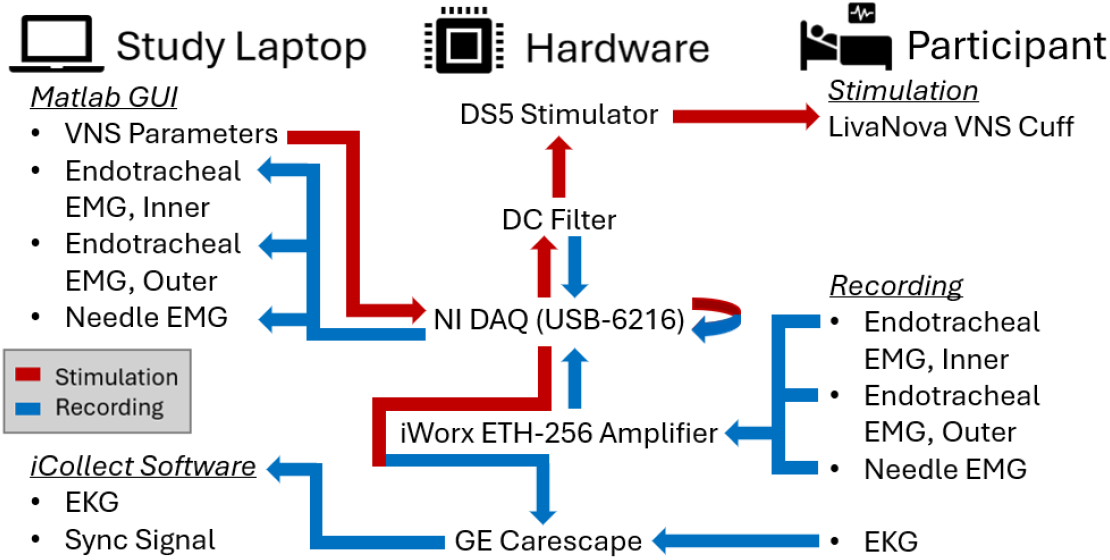
Diagram of the instrumentation setup and signal flow.

The three differential EMG signals—inner endotracheal, outer endotracheal, and needle (Figure 1)— were amplified (400-2000x) and filtered (0.3-2000 Hz) with low-noise voltage amplifiers (ETH-256, iWorx Systems Inc.; Dover, NH). The recording electrodes contralateral to stimulation (right side in all cases, red in Figure 1) were connected to the noninverting input of the recording amplifiers for EMG signals. The EMG signals were then digitized by the NI-DAQ at a sampling frequency of 50 kHz and recorded in MATLAB.

The EKG signal was recorded by a patient monitor (Carescape B650; General Electric; Boston, MA) using S/5 iCollect software (v5.0, General Electric; Boston, MA) at a sampling frequency of 100 Hz. To synchronize the EKG and stimulation signals, we connected the voltage-controlled stimulation output from the NI-DAQ to the Carescape monitor using a custom solid-state circuit (Supplement III: Figure 11).

### 3.3 Stimulation Trials

The experimental design is summarized in Table 1 and Figure 3. This protocol was delivered to 7 of 10 participants (P4 to P10). The first three participants (P1 to P3) had an earlier version of the protocol (Supplement IV, Supplement V), which informed protocol revisions to capture better the physiological (EMG and HR) responses, including additional lower stimulation amplitudes and fewer pulsewidths. The data from P1 to P3 were analyzed for trials with matched pulsewidths to P4 to P10.

**Table 1.** Experimental design of stimulation trials. The order of block delivery is diagrammed in Figure 3. All stimulation was delivered using symmetric biphasic rectangular pulses at 20 Hz. The “TEST” block serves to evaluate participant sensitivity to stimulation for setting a custom charge limit. The “CLIN” block uses the participant’s clinical settings. “CTRL” blocks are used interleaved with other blocks to assess biological drift as well as polarity effects. “LOW AMP” blocks (A, B, C) use lower stimulation amplitudes and shorter ON/OFF times aiming to capture the laryngeal EMG recruitment curves. “HIGH AMP” blocks (AA, BB, CC) use higher stimulation amplitudes and longer ON/OFF times aiming to capture changes in HR.

| Type | Block | Pulsewidth ($\mu\text{s}/\text{phase}$) | Amplitude (mA) | ON Time (s) | OFF Time (s) | # Pulses per ON Time |
| --- | --- | --- | --- | --- | --- | --- |
| TEST | TEST | 250 | 2, 4, 7 (not randomized) | 5 | 5 | 100 |
| CLIN | CLIN | 250 | Only for chronic participants; patient-specific clinical settings (Supplement VIII) | 5 | 5 | 100 |
| CTRL | X | 250 | 3.5, -3.5 | 15 | 15 | 300 |
| CTRL | XX | 250 | 3.5 | 15 | 15 | 300 |
| LOW AMP | A | 50 | 0.2, 0.4, 0.6, 0.8, 1, 1.2, 1.4, 1.6, 1.8, 2, 2.5, 3, 3.5 | 3 | 2 | 60 |
| LOW AMP | B | 250 | 0.1, 0.2, 0.3, 0.4, 0.5, 0.6, 0.7, 0.8, 0.9, 1, 1.5, 2, 2.5 | 3 | 2 | 60 |
| LOW AMP | C | 1000 | 0.05, 0.1, 0.15, 0.2, 0.25, 0.3, 0.35, 0.4, 0.45, 0.5 | 3 | 2 | 60 |
| HIGH AMP | AA | 50 | 3.5, 7.0, 10.0, 20.0, 25.0, 28.0 | 25 | 25 | 500 |
| HIGH AMP | BB | 250 | 1.0, 2.5, 3.5, 4.0, 6, 7 | 25 | 25 | 500 |
| HIGH AMP | CC | 1000 | 0.5, 1.0, 2.5, 3 | 25 | 25 | 500 |

**Figure 3.**
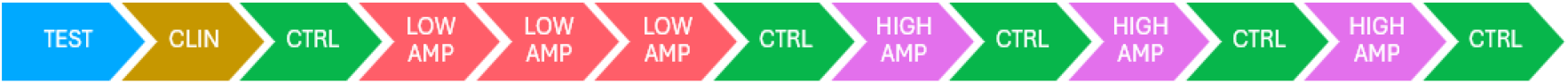
Experimental block flow diagram. Table 1 defines the block types. Example block order: X, B, A, C, XX, CC, XX, BB, XX, AA, X. See Supplement V for blocks and trials for each participant.

We delivered biphasic rectangular symmetric pulses at 20 Hz, with pulsewidths of 50, 250, and 1000 µs/phase over a range of amplitudes. We defined a “trial” as a given delivery of stimulation, with a specific pulsewidth, stimulation amplitude, ON time, and OFF time. We defined a “block” as a series of trials with fixed pulsewidth, ON time, and OFF time, and a range of stimulation amplitudes. In each block, we first recorded an OFF time, then alternated ON and OFF times for each trial, thus ending with an OFF time. We evaluated five types of blocks: TEST, CLIN (clinical parameter settings), CTRL (control), LOW AMP (lower stimulation amplitudes), and HIGH AMP (higher stimulation amplitudes). Except for the TEST blocks, the order of the stimulation amplitudes was randomized in each block for each participant. EMG and EKG signals were recorded continuously.

We limited the charge per phase to 3.5 µC. This limit was determined by the maximum settings available on the FDA-approved LivaNova VNS device: 1000 µs pulsewidth and 3.5 mA. Supplement VI: Charge Calculations provides calculations for the maximum allowable amplitude for each pulsewidth.

For five participants (P6 to P10), we first delivered a TEST block, stimulating at 2, 4, and 7 mA in ascending order with a pulsewidth of 250 µs/phase. The amplitudes in the TEST block were not randomized but rather served to evaluate patient sensitivity to stimulation. As shown in the decision tree of Supplement VII, if a trial in the TEST block resulted in a HR outside of the defined acceptable range (50-120 bpm), the stimulation amplitude was reduced by 1 mA and tested again until a limit which did not evoke unacceptable changes in HR was identified. If the HR was not within the acceptable range in response to 0.5 mA, we did not proceed with additional trials. We then used the patient-specific charge limit to exclude planned trials that exceeded the bound (Supplement VI: Charge Ratio Data).

After the TEST block, for chronic participants, we delivered the CLIN block (n = 4; P6 to P9; chronic participants P1 and P3 received the earlier version of the protocol that did not include the CLIN block; CLIN block was tested later in P8 (Supplement V)), in which we recorded responses to their clinically programmed pulsewidth and amplitude.

We then delivered LOW AMP blocks with pulsewidths of 50 (block A), 250 (block B), and 1000 (block C) µs/phase using stimulation amplitudes up to 3.5, 2.5, and 0.5 mA, respectively. Each trial was delivered for 3 s ON (60 pulses) and 2 s OFF. The low amplitudes and short durations of these trials were designed to measure the laryngeal EMG recruitment curves. The order of the three LOW AMP blocks was randomized for each participant.

Similarly, we delivered HIGH AMP blocks with pulsewidths of 50 (block AA), 250 (block BB) and 1000 (block CC) µs/phase using stimulation amplitudes up to 28, 7, and 3 mA, respectively. Each trial was delivered for 25 s ON (500 pulses) and 25 s OFF. The higher amplitudes and longer durations of these trials were designed to measure the effects of VNS on HR; the longer latency of HR change is not well-captured during the shorter LOW AMP blocks. The order of the three HIGH AMP blocks was randomized for each participant.

We delivered a CTRL block (block XX) after the CLIN block, after the three LOW AMP blocks, and after each HIGH AMP block (Figure 3) using 3.5 mA and 250 µs/phase. If the CTRL block charge was excluded due to a HR change outside of allowable bounds—as in P4 (Supplement V)—we reduced the amplitude to 2 mA. For some participants (n = 6; P4, P5, P6, P7, P9, P10), we also evaluated −3.5 mA—in a random order with the trial at +3.5 mA—to evaluate the effect of stimulation polarity (block X).

If the HR went beyond the bounds of 50-120 bpm during stimulation, as reported by the Carescape monitor, the stimulator was immediately manually deactivated. We then excluded any remaining trials that had higher charge—scaled depending on the pulsewidth (Supplement VI)—than the trial that caused that HR change. The entire protocol included 63 trials which required a total of 25 minutes, plus 3 minutes to connect and disconnect the research equipment. If there was insufficient time to deliver all trials, the planned protocol was followed until completion with one exception: if there was insufficient time to complete a HIGH AMP block (3.75 to 5.5 min per block), we skipped the HIGH AMP block and finished with a CTRL block (~1 min) if time allowed. All blocks and trials executed for each participant are provided in Supplement V.

### 3.4 Data Analysis

We analyzed the EMG and HR time series in MATLAB R2022b.

#### 3.4.1 Laryngeal EMG

We divided each EMG time series by the appropriate amplifier gain for the trials in the CLIN, CTRL, and LOW AMP blocks. For the endotracheal EMG channels, we observed low frequency responses during stimulation that were related to stimulation onset/offset and to stimulation amplitude but not related to the individual stimulation pulses (Supplement IX: Figure 14 and Figure 15). Therefore, to analyze the pulse-specific muscle responses, we applied a digital high-pass filter (FIR, Kaiser Window method) to the endotracheal EMG time series for each block using MATLAB’s “highpass” function and a corner frequency of 5 Hz (Supplement IX: Figure 16).

In one participant (P7), there was excessive powerline noise in the needle EMG recording. We applied a notch filter to each block time series (IIR, 2^nd^ order, Butterworth) with corner frequencies 55 and 65 Hz. We used MATLAB’s “filtfilt” function for zero-phase digital filtering.

To identify the time stamp of each stimulus pulse, we calculated the derivative of the recorded voltage pulse train output from the NI-DAQ and identified the peaks as the midpoint of each stimulus pulse. We then disregarded the first 3 ms after each pulse midpoint to exclude the stimulation artifact. We applied MATLAB’s “detrend” function to each trial to remove underlying baseline drift. For each of the three EMG channels and each trial, we calculated the stimulus-triggered median of 10 stimulation pulses (Supplement XI).

We calculated the area under the rectified curve (AUC) of the stimulus-triggered median of each trial using MATLAB’s “trapz” function for t = 5-24 ms from the midpoint of the stimulus pulse. We subtracted the noise floor from each calculated AUC; we calculated the noise floor as the AUC during the 1 ms preceding the first trial in each block, scaled by the duration of the signal window (19 ms). For each participant, recording channel and stimulation block, we normalized all denoised AUC values by the maximal AUC in that block (AUC_norm,_ Supplement XII). We excluded two trials from P1 with secondary muscle recruitment at high stimulation amplitudes (8.75 and 10 mA at 250 µs; Supplement XIII).

We fit the AUC_norm_ versus stimulation amplitude recruitment curve for each participant, recording channel, and block with a sigmoidal function using MATLAB’s “fit” function:

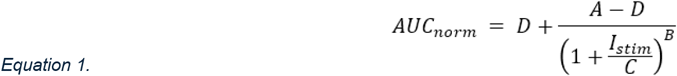

where A is the minimum value (fixed at 0), B describes the steepness of the curve, C is the inflection point, and D is the maximal value (fixed at 1). We defined the threshold for EMG activation as the stimulation amplitude required to evoke 50% of maximal response in the sigmoid fit, i.e., AUC_norm_(I_th50_) = 0.5.

For each recording channel, we fit the EMG I_th50_ thresholds for all participants across pulsewidths to a strength-duration curve using MATLAB’s “fit” function:

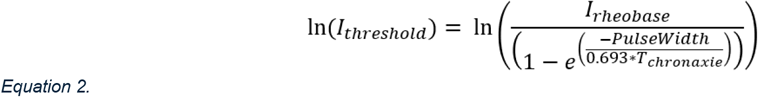

#### 3.4.2 Heart Rate

We used MATLAB’s “findpeaks” function to identify the peaks in the QRS waves in the EKG traces from which we calculated the HR using the R-R interval. We smoothed the HR time series by using a five-beat sliding window mean of HR, advanced in increments of one beat. Using the smoothed HR time series, we labeled each HIGH AMP, CLIN, and CTRL trial as having a bradycardic change in HR during stimulation, a tachycardic change in HR after stimulation, or no change in HR (Supplement XIV). This pre-processing strategy served to avoid quantifying the change in HR from trials with transient changes in HR that were unrelated to the trial stimulation. Trials were identified as having a bradycardic change by:

1. HR slope at onset of stimulation of <-1.5 bpm/s between the start of stimulation and the minimum HR during stimulation, OR
2. a decrease of ≥3 bpm during the trial ON time compared to the instantaneous HR at the start of stimulation AND greater mean HR in the following OFF time compared to the mean HR during stimulation.

Trials were identified as having a tachycardic change in HR by an increase of ≥3 bpm over the first 5 s of the OFF period.

For the trials that were identified as having a bradycardic change in HR during stimulation, we calculated the percent change in HR using the median HR over 15 s at the start of the OFF time preceding stimulation compared to the minimum HR during stimulation:

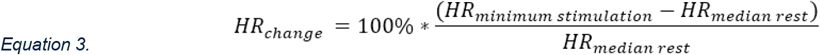

We only used the first 15 s of rest time to match data available between different protocols (P1-3 vs P4-10). For each participant and pulsewidth, we calculated the threshold for a 10% decrease in HR during stimulation ON using linear interpolation between the two lowest sequential amplitudes straddling HR_change_ = −10% (Figure 5B).

**Figure 4.**
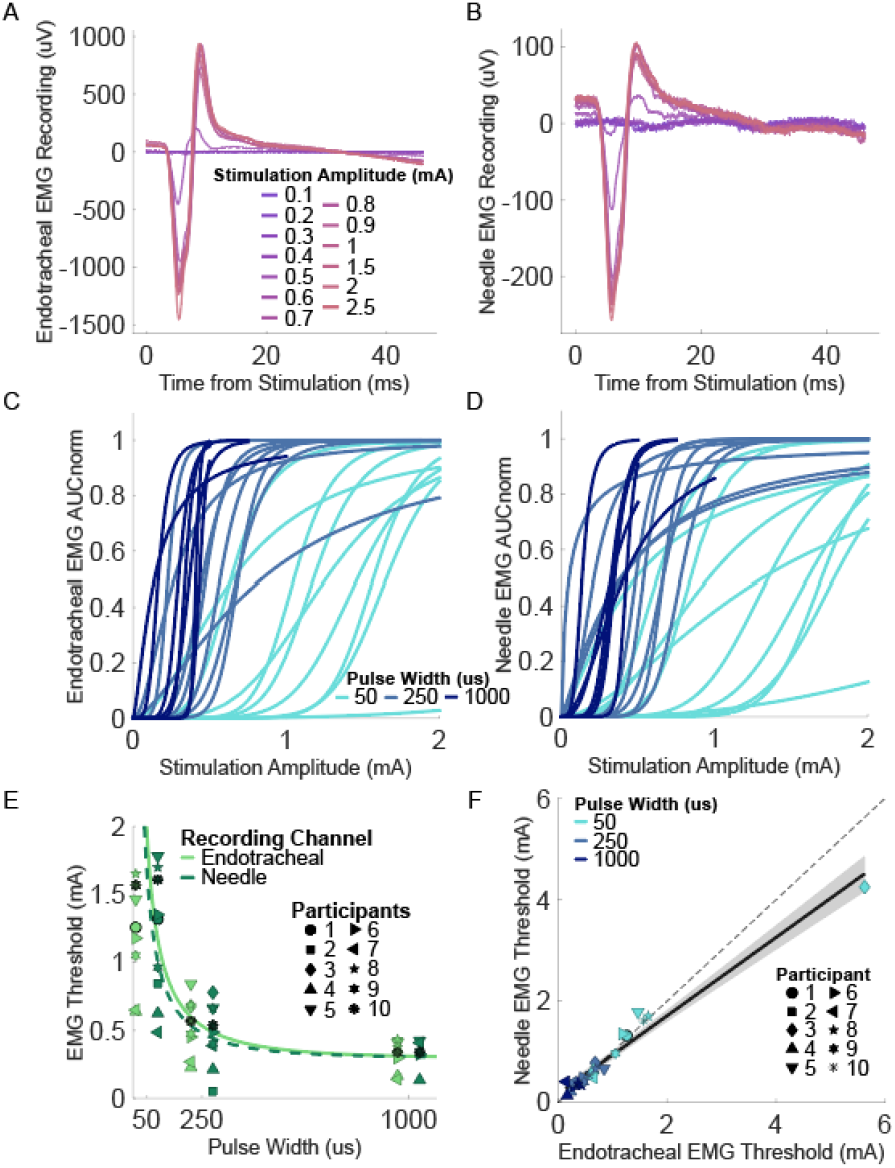
Laryngeal EMG thresholds for needle and endotracheal recordings. All trials from the endotracheal channel (A) and needle channel (B) in participant 10 during LOW AMP block B, pulsewidth = 250 µs. (C) Sigmoid fit of the normalized AUC of the endotracheal EMG for each participant and pulsewidth. (D) Sigmoid fit of the normalized AUC of the needle EMG for each participant and pulsewidth. (E) EMG thresholds across pulsewidths. P1 to P3 do not have a threshold for the 1000 µs pulsewidth because their protocol did not include enough trials to fit a sigmoid. (F) EMG thresholds for needle electrodes versus endotracheal electrodes. Black dashed line denotes 1:1 relationship. Black solid line denotes the linear regression. Grey shading denotes the 95% confidence interval (I_th_needle_ = 0.7725 mA/mA * I_th_endo_ + 0.1649 mA; R^2^ = 0.958).

**Figure 5.**
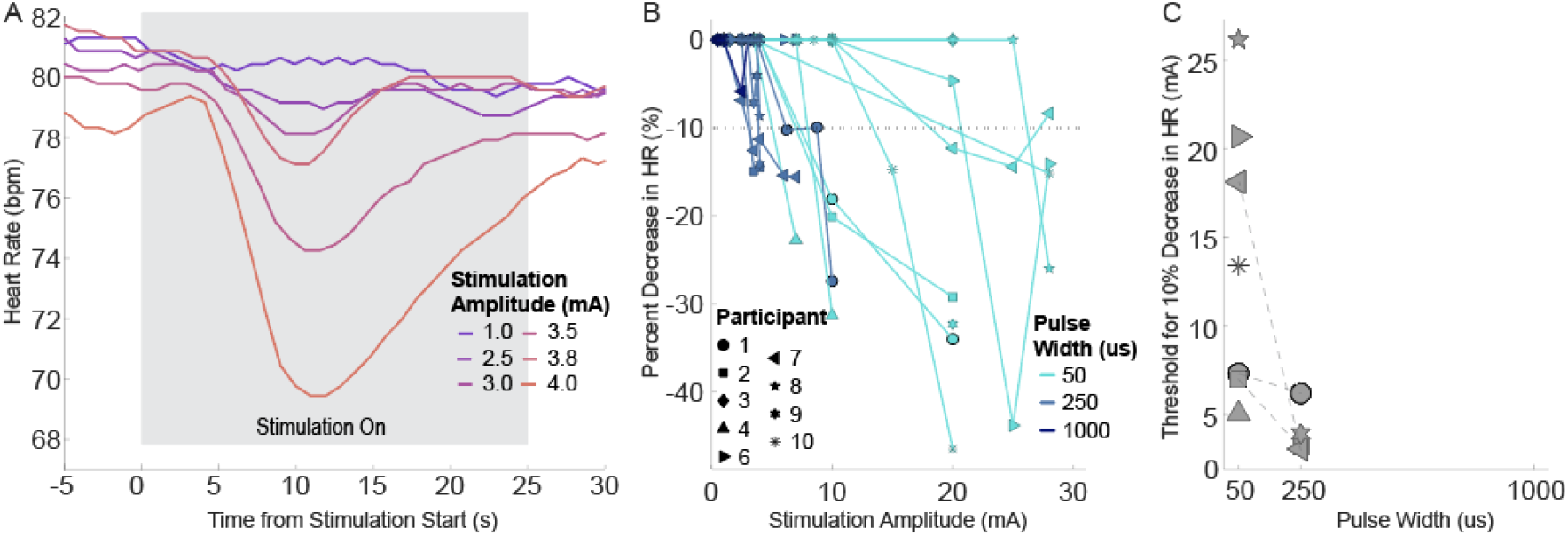
(A) Example heart rate (HR) traces during stimulation for HIGH AMP block BB (pulsewidth = 250 µs) for participant 9. (B) Decreases in HR across stimulation amplitudes and pulsewidths. (C) Thresholds for a 10% decrease in HR across pulsewidths. Lines connect thresholds from the same participant. No trials at 1000 µs evoked a change in HR.

Similarly, for trials that were identified as having a tachycardic change in HR after stimulation, we calculated the percent change in HR using the median HR over the first 15 s of the ON time compared to the maximum HR during the OFF time after stimulation; we determined the threshold for a 10% increase in HR using linear interpolation.

#### 3.4.3 Statistics

We conducted statistical analyses using JMP Pro (v17, JMP Statistical Discovery LLC, Cary, NC) and MATLAB R2023b. We evaluated the effects of four categorical fixed factors (EMG channel (endotracheal inner vs. endotracheal outer vs. needle), type of implant (acute or chronic), sex, and pulsewidth) and all interaction terms on EMG threshold in a linear mixed model in JMP, with participant as a random factor and zero intercept.

We used the concordance correlation coefficient function in MATLAB “f_CCC”^1^ to evaluate the similarity in EMG thresholds across recording channels.

We compared thresholds for bradycardia and tachycardia by ANOVA in MATLAB using the “anovan” function, with a full factorial of pulsewidth and type of threshold (bradycardic vs. tachycardic). We used a paired t-test in MATLAB “ttest” to compare the effects of stimulation polarity (cathodic leading cranial vs. anodic leading cranial) on EMG latency, EMG amplitude, and minimum HR during stimulation. We conducted an unpaired t-test in MATLAB “ttest2” to compare electrode impedance in acute vs. chronic participants.

We fit a linear regression to the HR thresholds vs. EMG thresholds and tested significance using the linear model function in MATLAB “fitlm”. We compared the regressions for acute vs. chronic participants using ANCOVA in MATLAB with the “aoctool”; the interaction term between the grouping variable (implant type) and the independent variable in the regressions (EMG threshold) determines whether the regression slopes are significantly different.

## 4. Results

We recorded laryngeal EMG and EKG in participants undergoing surgery for an implanted VNS device to treat epilepsy, either an IPG replacement (“chronic”) or new (“acute”) implant. Intraoperative data collection lasted 30 minutes, and we analyzed the thresholds to evoke EMGs and changes in HR.

### 4.1 Thresholds for Laryngeal EMG Responses

We recorded laryngeal EMG responses to stimulation via three differential EMG recording channels (Figure 1): endotracheal inner, endotracheal outer (Figure 4A), and needle (Figure 4B). Given the proximity of the inner and outer endotracheal channels, the similarity of their recorded responses, and the comparable or larger signal amplitude from “outer” compared to “inner” (10/10 participants, Supplement XII: Figure 20, Figure 21), all results compare only endotracheal outer (henceforth labeled “endotracheal”) and needle EMG recordings.

Longer pulsewidth resulted in lower EMG thresholds (Figure 4C-D, Supplement XV). The median and range of EMG thresholds (I_th50_) were 1.32 (0.49-5.63), 0.49 (0.05-0.84), 0.34 (0.13-0.43) mA for pulsewidths of 50, 250, and 1000 µs/phase, respectively (needle and endotracheal channels combined; n = 10, 9, and 7 participants for 50, 250, and 1000 µs/phase, respectively). The rheobase and chronaxie were 0.301/0.298 mA and 253/305 µs for needle/endotracheal, respectively (Figure 4E).

We did not detect a difference between EMG thresholds for 50% recruitment with endotracheal versus needle electrodes (Figure 4F). The Pearson correlation coefficient was high (R^2^ = 0.958; slope was close to 1 (0.77), p < 0.0001), and the concordance correlation coefficient showed strong similarity (p_c_ = 0.9511).

Within each channel, the maximum evoked EMG response was comparable across pulsewidths (Supplement XII: Figure 19, Figure 20, Figure 21). We observed some drift in EMG amplitude but not latency across the CTRL blocks, especially in P3, P6, and P7 (Supplement XVI: Figure 25, Figure 26). In P3, the first CTRL block elicited a larger response in the needle channel, but in following tests, the amplitude was consistent, suggesting mechanical settling of the needle positioning; conversely, the amplitude of the endotracheal channel was consistent in all CTRL blocks for P3 (Supplement XVI: Figure 27). For P6 and P7, the change in CTRL EMG amplitude primarily occurred after the LOW AMP, EMG-focused trials.

### 4.2 Thresholds for Bradycardia and Tachycardia Responses

VNS evoked a ≥10% decrease in HR in 8/9, 4/9, and 0/9 participants at pulsewidths of 50, 250, 1000 µs/phase, respectively. HR decreased in response to stimulation onset and began to recover before stimulation cessation (Figure 5A). Three participants (P4, P9, P10) exhibited stimulation-evoked asystole (Supplement V); normal sinus rhythm immediately resumed upon cessation of stimulation in all cases. The median and range of thresholds to evoke a 10% decrease in HR were 13.39 (5.04 – 26.19) mA at 50 µs/phase (n = 7) and 3.53 (3.05 – 6.19) mA at 250 µs/phase (n = 4) (Figure 5B, C).

We observed “rebound” tachycardia following cessation of stimulation in 5/9 participants (Figure 6A). We did not detect differences in tachycardia threshold and HR threshold (Figure 6B; ANOVA, p = 0.07).

**Figure 6.**
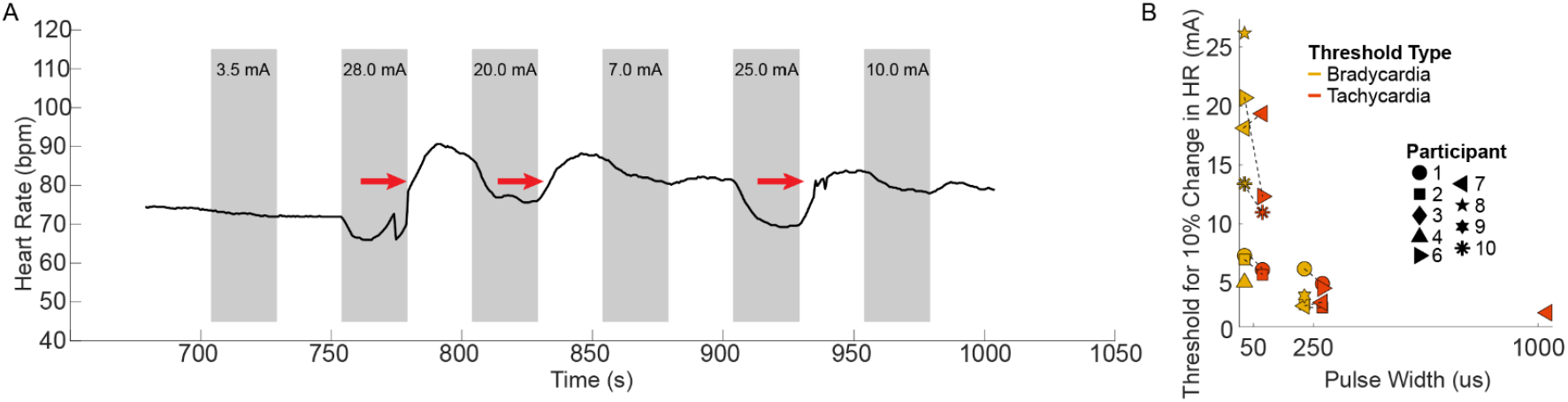
(A) Example heart rate (HR) time series from participant 7 during HIGH AMP block AA, pulsewidth 50 µs. The grey blocks denote stimulation ON. Tachycardia (red arrows) occurred after stimulation at 28, 20, and 25 mA. (B) Comparison of thresholds to evoke 10% bradycardia during stimulation and 10% tachycardia after stimulation.

### 4.3 Correlation Between EMG and HR Thresholds

We quantified the relationship between needle EMG and HR thresholds to determine whether laryngeal EMG—which can be evoked at low stimulation amplitudes—can serve as a predictor for stimulation amplitudes required to activate small, myelinated fibers. For participants (n = 8) and pulsewidths for which both EMG and HR thresholds were available, the HR thresholds were 9.02x (5.55 to 63.38x) higher than the needle EMG thresholds. EMG and HR thresholds were positively correlated (R^2^ = 0.477) with a slope of 9.93 mA/mA (p = 0.01) (Figure 7A). Figure 7B shows linear regressions of HR vs. EMG thresholds separated by acute and chronic participants. There is a slightly higher HR threshold relative to EMG threshold in chronic participants (slope of 11.28 in chronic vs. 6.73 in acute), but we did not detect a significant difference in the slopes of the correlations of EMG and HR threshold between acute and chronic participants (ANCOVA, p = 0.5788).

**Figure 7.**
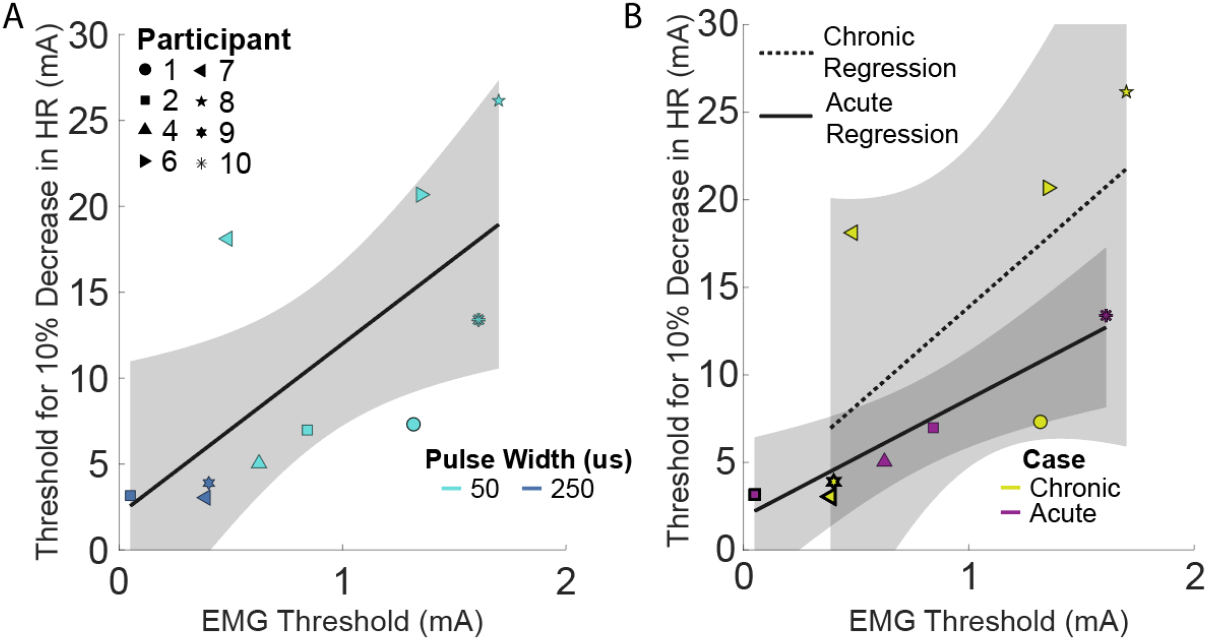
Regression of needle EMG thresholds and HR thresholds for all data color-coded by PW (A) and by case type (B). The black lines denote linear regressions, and the grey shadings denote 95% confidence intervals. (A) I_th_HR_ = 9.93 mA/mA * I_th_needle_ + 2.06 mA; R^2^: 0.477, p = 0.027. (B) Chronic: I_th_HR_chronic_ = 11.28 mA/mA*I_th_needle_chronic_ + 2.58 mA; R^2^: 0.62; p = 0.03. Acute: I_th_HR_acute_ = 6.73 mA/mA *I_th_needle_acute_ + 1.88 mA; R^2^:0.989; p = 0.005. Thick marker outline denotes 250 µs thresholds.

Neither EMG nor HR thresholds correlated with acute vs. chronic, sex, or electrode impedance (Supplement XVII); we did not conduct statistical analyses due to the small sample size. Electrode impedances were higher in chronic participants than acute participants (p = 0.031, unpaired t-test, Supplement XVIII). Because all our chronic participants were self-reported responders, we were unable to compare thresholds for laryngeal EMG and HR between responders and non-responders.

### 4.4 Responses to Clinically Programmed Amplitudes

Clinical parameters were set to 250 µs and 20 Hz for all 6 chronic participants (Supplement VIII). Clinical stimulation amplitudes were higher than EMG thresholds and lower than HR thresholds at 250 µs. Specifically, the median clinical amplitude was 2 mA (range of 1.75 to 3 mA); the EMG threshold was ~4x smaller (0.49 mA; 0.05 to 0.84 mA, Figure 8B) and the HR threshold was ~2x larger (3.53 mA; 3.05 to 6.19 mA, Figure 8C). For each chronic participant, the clinical amplitude and pulsewidth combination is on the maximal plateau of the EMG recruitment sigmoid (Figure 8A). We did not observe changes in HR with stimulation at clinical settings in the CLIN blocks.

**Figure 8.**
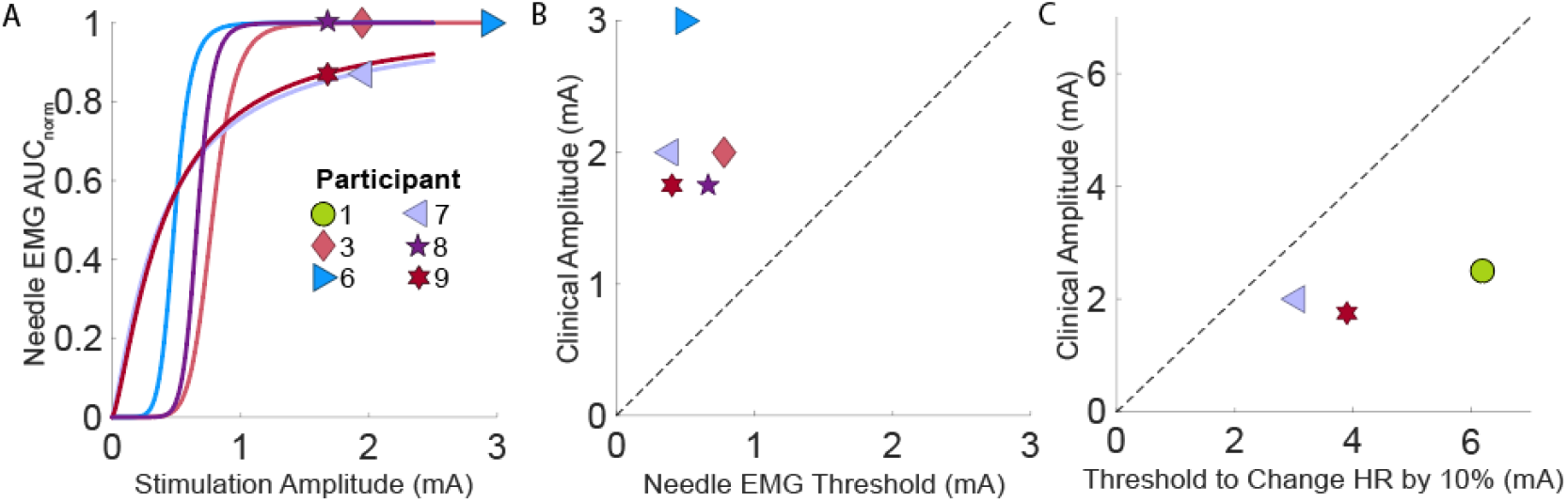
(A) Each participant’s clinical amplitude marked on their recruitment sigmoid for needle EMG recordings at their clinical pulsewidth of 250 µs/phase. Clinical amplitudes compared to (B) needle EMG and (C) heart rate (HR) thresholds at 250 µs. Dashed lines denote a 1:1 relationship.

Nerve fibers within the VNS cuff include those that form the recurrent laryngeal nerve. Activation of these fibers causes contractions of the laryngeal muscles, which causes the common side effects of VNS. All chronic participants reported voice changes as a side effect of VNS (n = 6); coughing (n = 4) and hoarseness (n = 4) were the next most common side effects (Supplement I: Table 3).

### 4.5 Effects of Stimulation Polarity

VNS treatment for epilepsy and depression involves delivery of the cathode-leading pulse to the cranial contact, whereas VNS for heart failure involves delivery of the cathode-leading pulse to the caudal contact. The premise is that the anode-leading waveform will block propagation (Ahmed et al., 2020), thus leading to unidirectional activity. However, computational modeling suggests that anodic block does not occur in human VNS (Musselman et al., 2023b). We did not detect an effect of polarity on EMG amplitude (Figure 9A; p = 0.89, paired t-test) or latency (Figure 9B; p = 0.85, paired t-test). Some changes in HR were observed during both polarities of stimulation, but we did not observe a difference between stimulation polarities (Figure 9C; p = 0.67, paired t-test). However, not all participants had a measurable change in HR, so we also evaluated the minimum HR during stimulation. As with the EMG and percent change in HR, we did not observe an effect of polarity on minimum HR during stimulation (p = 0.56, paired t-test, data not shown).

**Figure 9.**
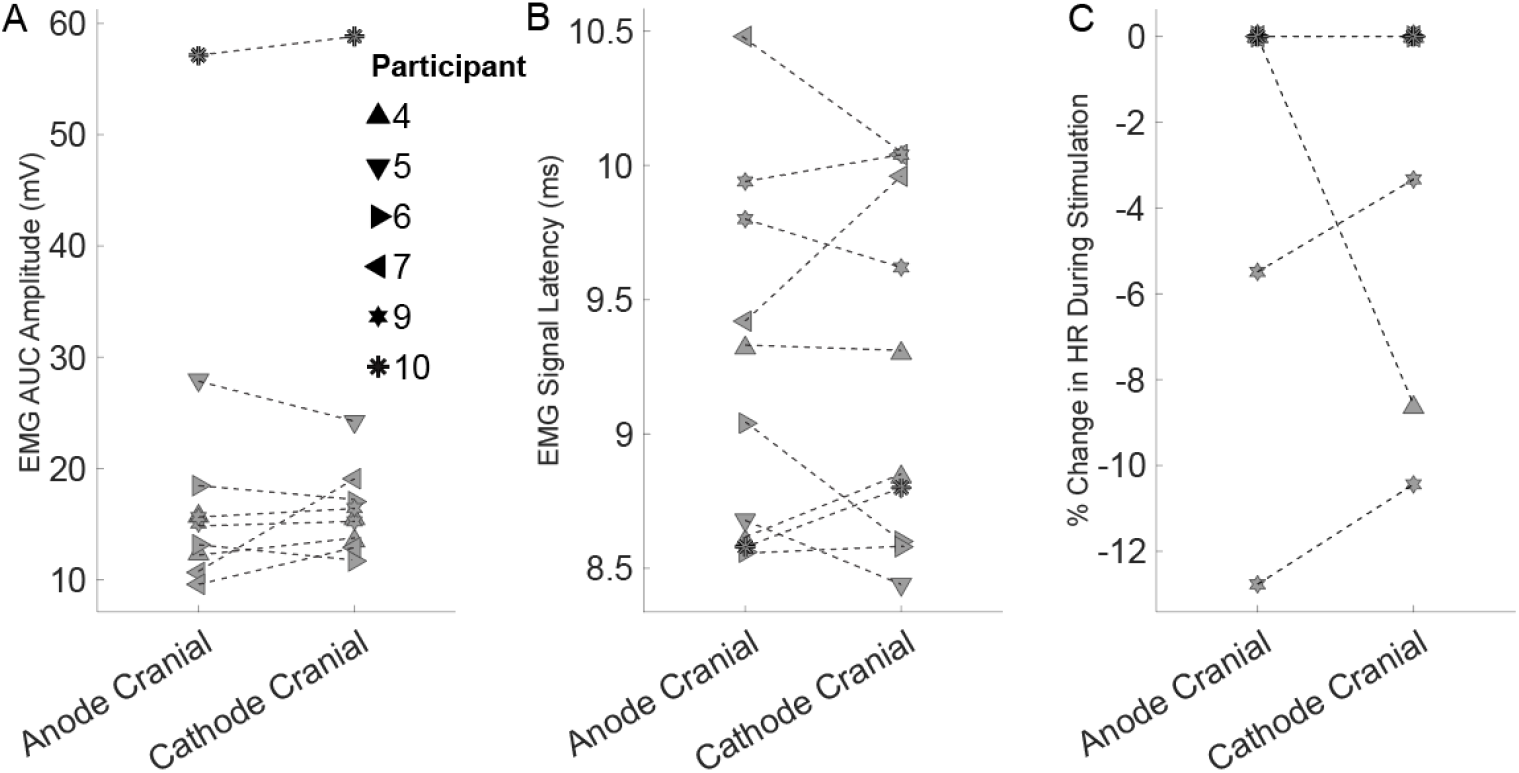
Effect of polarity on EMG and HR responses. (A) AUC of the needle EMG response, (B) latency of maximum peak of the needle EMG signals, and (C) percent change in HR during stimulation.

## 5. Discussion

We quantified physiological responses to VNS including laryngeal muscle activation and changes in HR. These data expand our understanding of the activation thresholds for different nerve fiber types in human VNS. Prior literature lacks responses to systematic sweeps of stimulation amplitudes and pulsewidths and VNS thresholds for evoking bradycardia in humans. In addition to informing the recruitment of side effects and therapeutic settings, such data are necessary for validating computational models of VNS (Musselman et al., 2023b).

### 5.1 Laryngeal EMG Responses

We conducted laryngeal EMG recordings during VNS using simultaneous invasive (endotracheal) and non-invasive (needle) approaches. The endotracheal electrodes are closer to the signal source (laryngeal muscles: posterior and lateral cricoarytenoid, cricothyroid, arytenoid, and thyroarytenoid), resulting in higher signal-to-noise ratio (SNR), whereas needle or surface electrodes can be used in clinic, with awake participants, without anesthesia or paralytic. The non-invasive needle electrodes enabled equivalent detection of EMG thresholds to the invasive endotracheal electrodes, and this provides important validation of the non-invasive approach; prior studies used surface electrodes to record laryngeal EMG in awake patients as a proxy for vagal activation (Bouckaert et al., 2022; Vespa et al., 2019). Indeed, preclinical VNS studies demonstrated correlation between A-fiber activation and laryngeal EMG response in dogs (Castoro et al., 2011), rats (Chang et al., 2020), and pigs (Blanz et al., 2023). Our EMG thresholds were comparable to the results in prior human subjects studies, all for a 250 µs pulsewidth: 0.49 mA in our study (median; range of 0.05-0.84 mA) vs. 0.3-0.75 mA in literature (Bouckaert et al., 2022; Vespa et al., 2019).

During LOW AMP blocks and especially HIGH AMP blocks, we observed low frequency activity in the endotracheal EMG recordings during stimulation (onset response) and after stimulation (offset response) (Supplement IX). This low frequency activity decreased in amplitude with time after stimulation ON or OFF, with a period of ~0.5 to 3 s; its amplitude was near zero after ~1 to 6 s. The signal’s duration was longer with higher stimulation amplitude. The longest durations were measured in HIGH AMP blocks which were not included in EMG analysis. Given that this low frequency activity only appeared on the endotracheal EMG channels and not the needle EMG channels, it may reflect tonic tracheal contractions.

The first three participants had a different experimental design. The preliminary protocol informed the final protocol to help ensure that we captured the EMG recruitment curve and that stimulation amplitudes were sufficiently high to evoke changes in HR. The preliminary protocol only included two trials at 1000 µs, and thus the EMG responses could not be fit to a sigmoid. Additionally, trials were not comprehensive of low amplitudes (<0.5 mA) at 250 and 1000 µs which further increased the challenge of fitting recruitment curves.

At high amplitudes, we occasionally observed recruitment of secondary muscle groups in the EMG channels (Supplement XIII). This recruitment may be due to current leakage that activated other neural pathways outside of the cuff electrode (Nicolai et al., 2020). This was defined as an obvious change in waveform shape and primary peak latency and sometimes polarity (positive or negative). This most often occurred in the HIGH AMP blocks and so did not complicate the LOW AMP block EMG analyses for P4-P10. However, in P1, where all amplitudes for a given pulsewidth were tested in a single block, we excluded responses to 8.75 and 10 mA in our EMG analysis for this reason.

In one participant, P7, there was a low SNR in the endotracheal inner and needle EMG channels. The contaminating noise had a large 60 Hz component, and the notch filter we implemented did not completely remove the artifact. This made measurement of the EMG amplitude difficult and resulted in noisy data with a weaker sigmoidal fit. There was no notable difference in this case which might have caused this noise.

### 5.2 Heart Rate Responses

We demonstrated that bradycardia can be evoked by VNS in humans. This has been an elusive effect in literature, with reports of a decrease in HR by ~4 bpm in a study of 59 patients (half left and half right VNS) (Premchand et al., 2014), case reports of extreme VNS-evoked bradycardia or asystole (HR < 35 bpm) (Åmark et al., 2007; Iriarte et al., 2009; Pascual, 2015; Shankar et al., 2013), or no bradycardia evoked in any patients (n = 6, n = 55) (Banzett et al., 1999; Zannad et al., 2015). Our thresholds for bradycardia were several times higher than the clinical VNS parameters for our participants: 3.5 mA vs. 2 mA at 250 µs.

We observed “rebound” tachycardia following cessation of stimulation for some trials, which has also been reported in preclinical studies (Deshmukh et al., 2025). Tachycardia occurred without prior bradycardia in some trials, and conversely, tachycardia was absent following some trials with bradycardia (Supplement XIX: Figure 30). The thresholds to evoke a 10% decrease in HR during stimulation and a 10% increase in HR after stimulation were not significantly different. We did not observe tachycardia during stimulation, which can occur at low amplitudes due to a reflex response following activation of afferent fibers (Ardell et al., 2017, 2015) or due to activation of sympathetic fibers within the vagus nerve (Seki et al., 2014); we may have observed the former effect if our “HIGH AMP” blocks that were HR-focused (with longer ON and OFF times) included lower stimulation amplitudes.

Quantification of stimulation-evoked changes in HR can be confounded by the time required for HR to return to baseline following preceding trials. “Rebound” tachycardia confounded measurement of bradycardia in some cases where baseline HR had not recovered in the 25 s allowed between trials. Additionally, in P1 to P3, a different block design was used, and the ON and OFF times were only 15 s; this was sufficient to quantify bradycardia, which peaked at 8.2 s (median; range of 4.3 to 14.8 s), but did not provide sufficient time to recover from rebound tachycardia. In future studies, we recommend longer OFF time between HIGH AMP trials. Our trial design prevents systematic analysis of recovery times, but it can be up to or exceeding one minute based on blocks where tachycardia occurred on the last trial. Conversely, the active stimulation time could be reduced from 25 s to 15 or 20 s.

P3 is the only participant that had no changes in HR during stimulation. This participant was given vasopressin to stabilize low blood pressure at the beginning of surgery. Vasopressin enhances the reflexive constriction of blood vessels (Ebert et al., 1986) which may have affected the HR response to VNS.

### 5.3 Relative Thresholds for EMG and HR Responses

The therapeutic effects of VNS for epilepsy are proposed to be mediated by activation of afferent fibers (Fraschini et al., 2013; Groves et al., 2005; Krahl et al., 1998) while dominant side effects are produced by activation of large diameter myelinated somatic efferent fibers (Kersing et al., 2002; Vespa et al., 2019). The clinical stimulation amplitudes in chronic participants were well above (~4x) the activation thresholds for laryngeal EMG but below the threshold to evoke bradycardia (~2x). Clinical waveforms are asymmetrical, with a passive recharge phase, whereas we used symmetric pulses, which may increase thresholds by ~1.3-2x (Gorman and Mortimer, 1983; van den Honert and Mortimer, 1979). Biphasic stimulation also enhances separation between thresholds for fibers of different diameters (Gorman and Mortimer, 1983) which may have caused a larger separation between laryngeal EMG threshold and HR threshold in our study. Similar ratios (~4x, 1-10x mA/mA mean and range) are reported for B to A fiber activation ratios in preclinical studies (Blanz et al., 2023; Castoro et al., 2011; Chang et al., 2020; McAllen et al., 2018; Thompson et al., 2025; Yoo et al., 2013). However, anatomy and functional organization differ between species (Jayaprakash et al., 2023; Kronsteiner et al., 2022; Pelot et al., 2020; Settell et al., 2020) which may explain why bradycardia is more often reported during VNS in animals. Additionally, these studies were conducted with animal subjects under anesthesia, which allows for much higher stimulation intensities, which in turn can evoke bradycardia.

Within participants, thresholds for EMG and HR were positively correlated (Figure 7): participants with higher EMG thresholds had higher HR thresholds. There is individual variability in thresholds, which points to the need for systematic approaches to stimulation parameter selection. In a computational study of human and animal VNS, activation thresholds vary widely between individuals of the same species; using the same or linearly scaled stimulation did not produce equivalent activation between species (Musselman et al., 2025). All chronic participants (n = 6) self-identified as VNS responders, with VNS improving their condition by reducing frequency and severity of seizures (Supplement I: Table 3). This may indicate that the small diameter afferent fibers that mediate therapeutic effects are of a larger diameter than the small diameter efferent fibers that mediate bradycardia. Alternatively, subthreshold responses may be therapeutic, or more fiber activation is required to produce bradycardia as compared to reducing seizure frequency or severity.

### 5.4 Unidirectional Activity Evoked by VNS

In VNS for drug resistant epilepsy, the cathodic-leading pulse is delivered to the cranial contact. When VNS was applied for heart failure, the cathode was placed caudally (Premchand et al., 2014). The orientation of the electrode has been suggested to create directional therapy by anodic block (Ahmed et al., 2020; Arle et al., 2016). However, modeling indicates that human VNS does not cause block at clinical amplitudes (Musselman et al., 2023b). Indeed, we found that polarity did not affect activation of laryngeal EMG or HR. This may indicate that the difference in polarity for treating epilepsy and depression (cathode cranial) compared to heart failure (cathode caudal) is unimportant for achieving therapeutic effects.

## 6. Conclusions

We measured VNS thresholds for laryngeal muscle activation and bradycardia in humans. Thresholds to evoke bradycardia are ~2x higher than clinically programmed amplitudes, indicating that the vagus nerve fibers that mediate epilepsy therapy may be of larger diameter than cardiac-projecting fibers responsible for stimulation-induced bradycardia. Alternatively, the therapeutic effect in refractory epilepsy may be evoked with less fiber activation than that required for bradycardia, or the therapeutic effect may be produced with subthreshold depolarization of these small diameter fibers. There was high concordance between needle and endotracheal channels of laryngeal EMG recordings. These data are useful for validating computational models of human VNS for device design and may be informative for programming clinical devices.

## Data Availability

All data produced in the present study are available upon reasonable request to the authors, at this time.

## 7. Acknowledgments

The authors would like to acknowledge funding from the Duke-Coulter Translational partnership. We would also like to thank the participants for their time and making this study possible. We thank the clinical staff, and clinical engineering at the Duke Medical Center for their support and cooperation in making this research possible. We thank Dr. Beiyu Liu and Dr. Shein-Chung Chow from the Duke Biostatistics Epidemiology and Research Design (BERD) team and Dr. Theodore Slotkin for consultation on the statistical analyses. We thank Princess Tara Zamani and Lucas Kaplan for support with intraoperative data collection. We thank Emma Lin and Zhecheng Dong for data analysis support. We thank Dr. Steve Schmidt for assistance with equipment setup and early study execution.

## Footnotes

1 robertpetermatthew/f_CCC: MATLAB code for computing Lin’s Concordance Correlation Coefficients including confidence intervals

