## Supplemental Data for "Intraoperative laryngeal muscle and heart rate responses to implanted vagus nerve stimulation"

### Supplement XIII

#### *Secondary Recruitment in Participant 1*

P1 showed some secondary recruitment at high amplitude stimulations in multiple channels (Figure 22). We identified this as secondary recruitment by the change in latency of the first peak as well as the change in waveform shape. We excluded these trials (8.75 and 10 mA at 250 µs pulsewidth) from our EMG analysis.

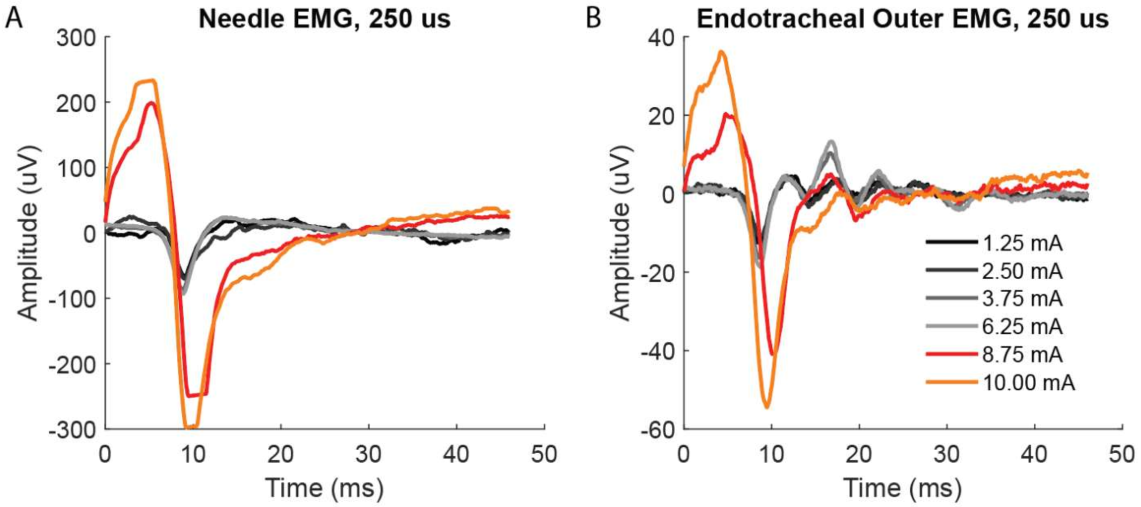


*Figure 22. Participant 1, block B100. Examples of secondary recruitment in response to 8.75 mA (red) and 10 mA (orange). The waveform shape as well as latency and direction of the first peak are different compared to the responses to lower stimulation amplitudes (grey), indicating secondary recruitment. These trials were excluded from sigmoidal fits.*

### Supplement XIV

#### *HR Change as Unrelated to Stimulation*

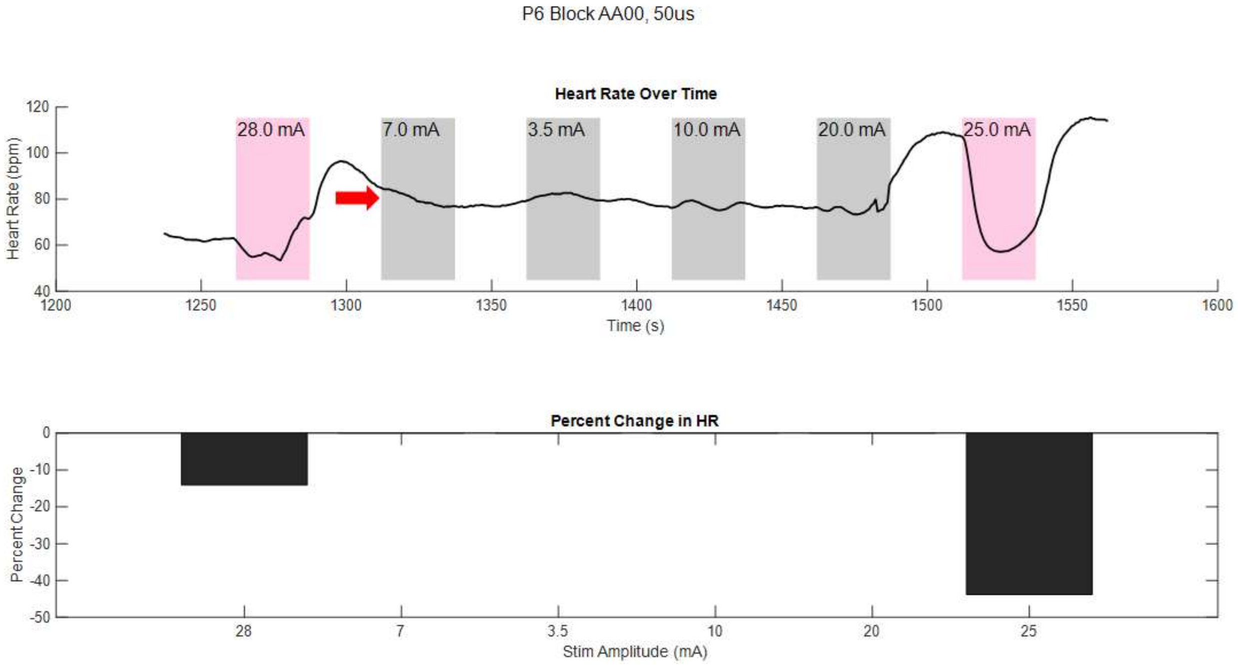


*Figure 23. Trial with stimulation-evoked bradycardia were identified using the rules described in the main text. This enabled exclusion of trials where the heart rate decreased, by not due to stimulation (e.g., red arrow, where heart rate decreased steadily during stimulation at 7 mA but without apparent relation to stimulation being on). Top: heart rate over time, stimulation is indicated by shaded boxes, pink = stimulation-evoked change in heart rate, grey = no change in heart rate detected. Bottom: percent change in heart rate.*

### Supplement XV

#### *EMG Recruitment Curves: All Recording Channels in All Participants*

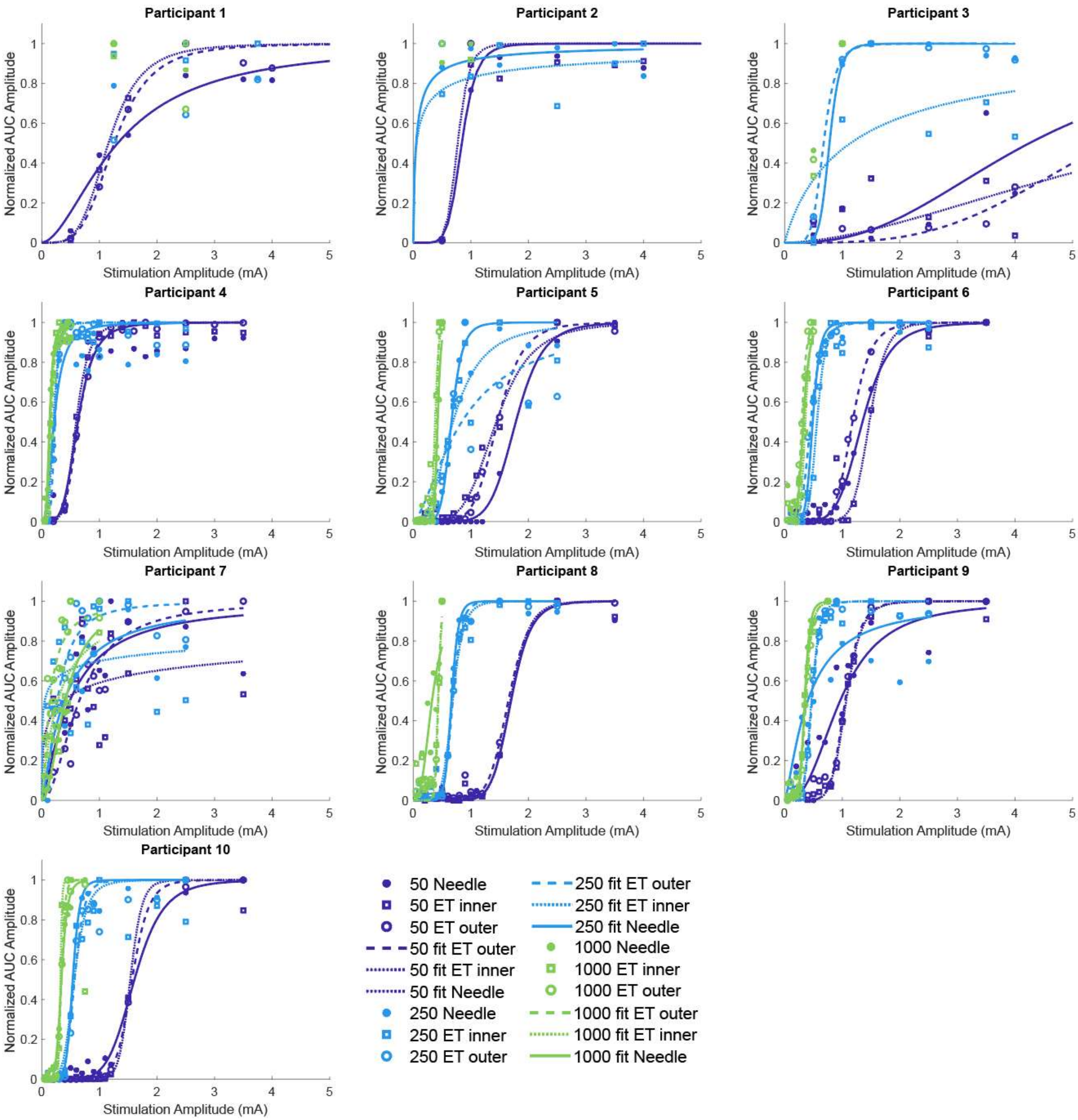


*Figure 24. All sigmoidal recruitment curves for each participant across pulsewidths.*

### Supplement XVI

#### *EMG Amplitude and Latency in CTRL blocks over the case*

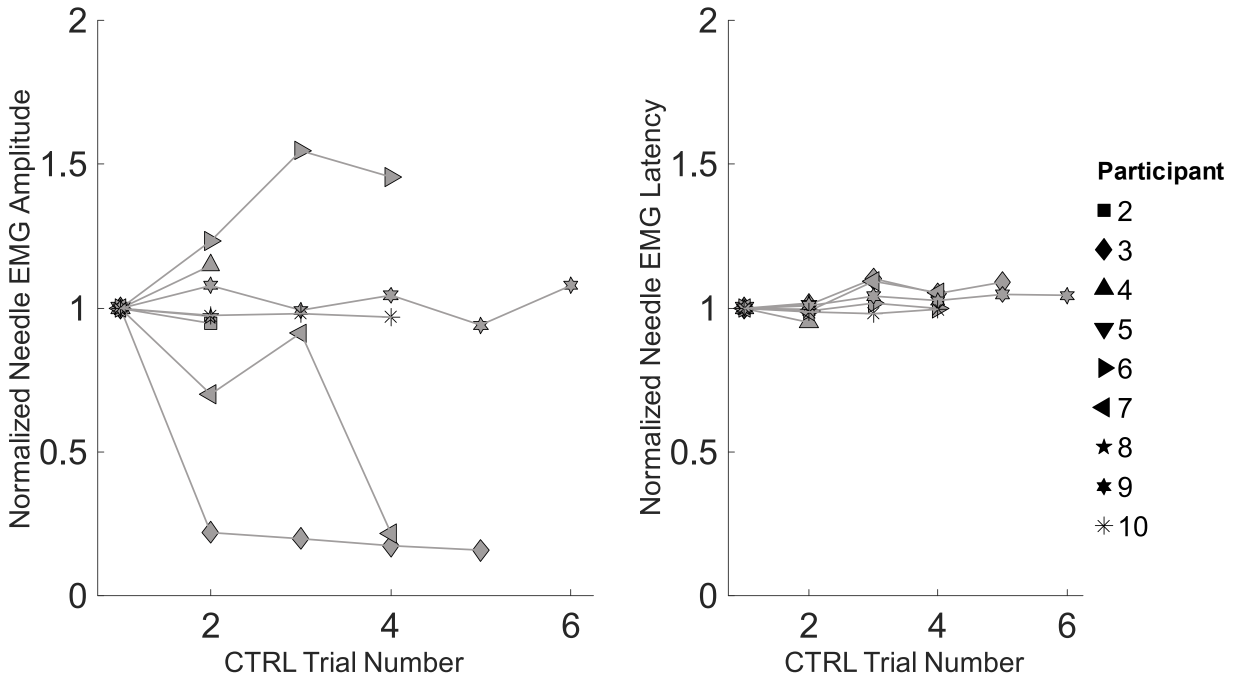


*Figure 25. Amplitude and latency of EMG recordings on needle EMG channel in CTRL blocks executed over the course of research time.*

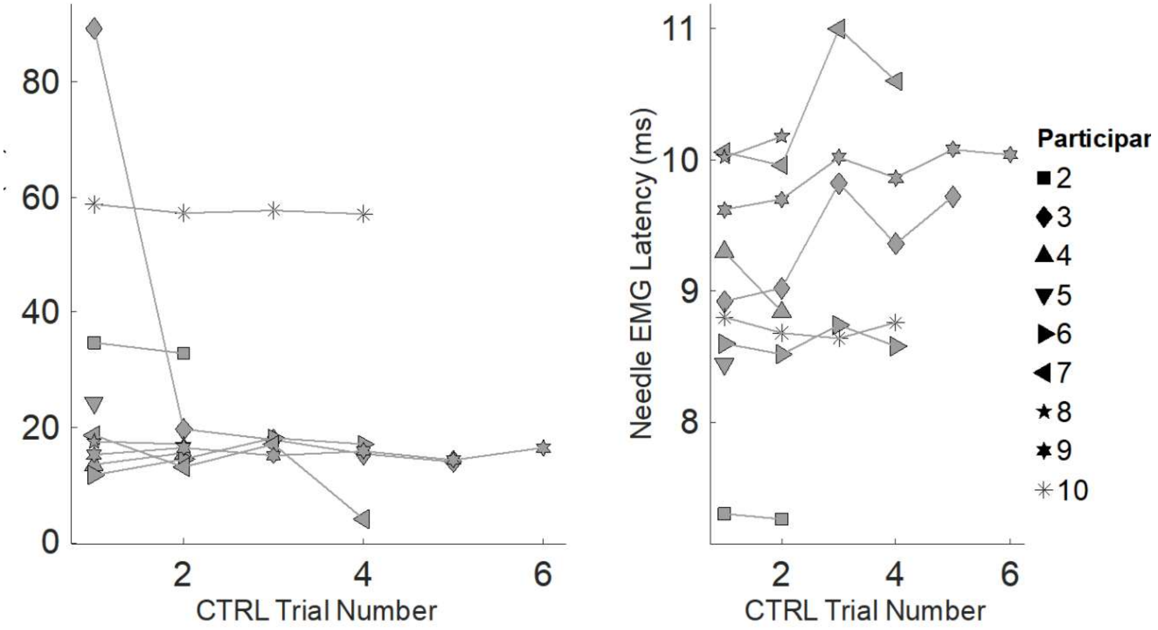


*Figure 26. Non-normalized version of Figure 25.*

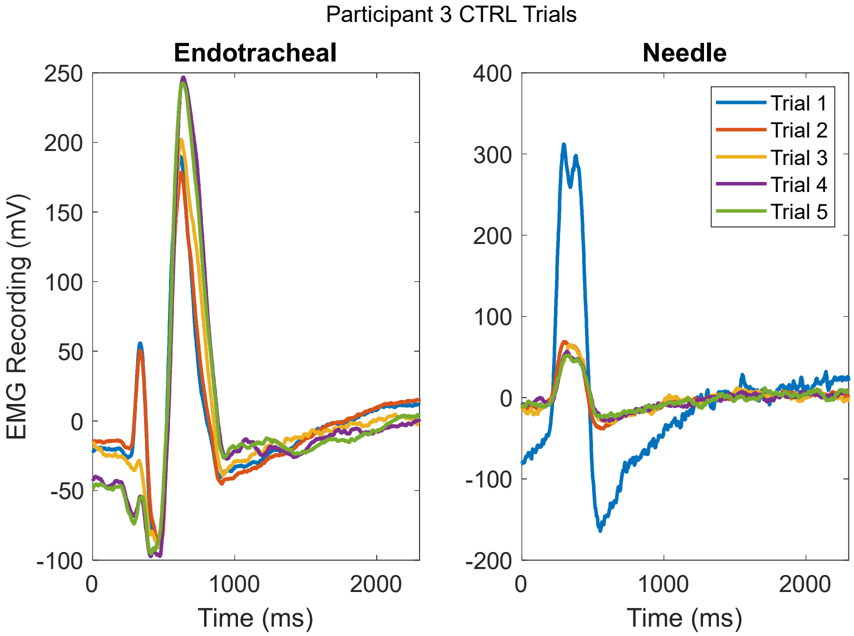


*Figure 27. Recorded response to CTRL block in P3 on endotracheal and needle EMG channels. Amplitude is higher in the first CTRL trial of the needle EMG but is stabilized in subsequent CTRL trials.*

### Supplement XVII

#### *EMG and HR Thresholds vs Patient Characteristics and Electrode Impedance*

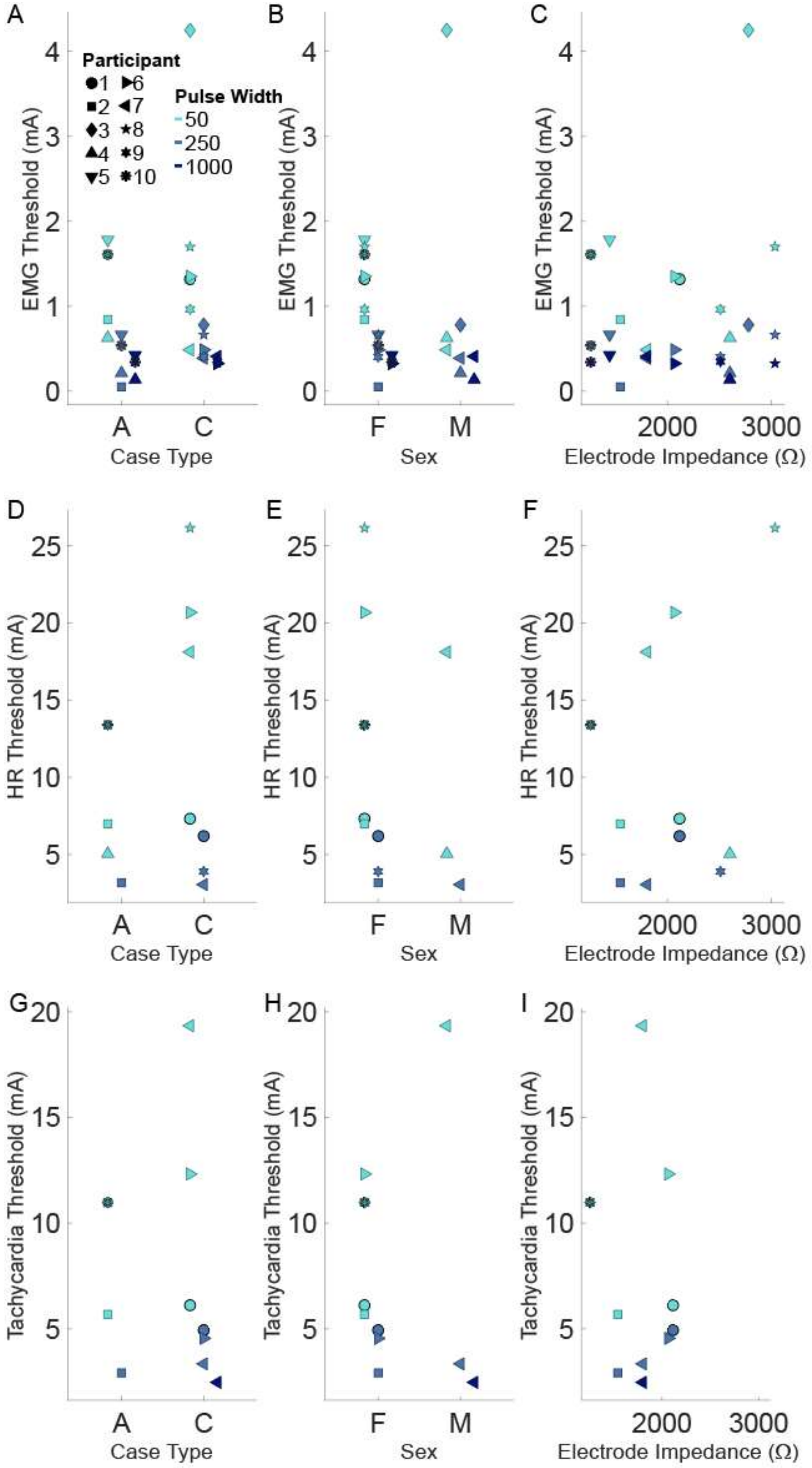


*Figure 28. Needle EMG threshold (A-C), bradycardia HR threshold (D-F), and tachycardia HR threshold (G-I) grouped by acute or chronic case (A,D,G), sex (B, E, H), and electrode impedance (C, F, I), color-coded by pulsewidth. Low N prevents statistical comparison.*

### Supplement XVIII

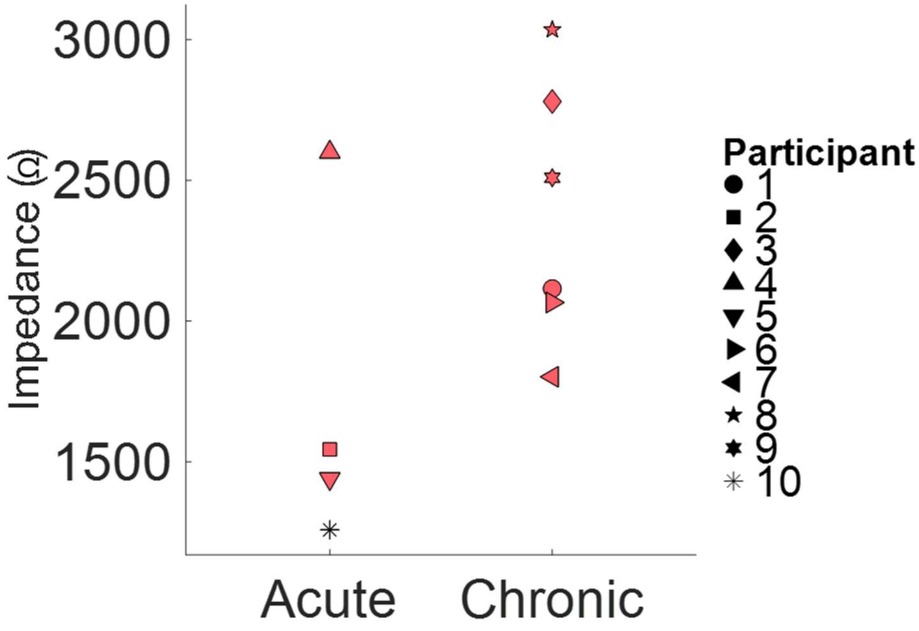


*Figure 29. Electrode impedance vs case type.*

### Supplement XIX

#### *Tachycardia Occurs with and without Preceding Bradycardia*

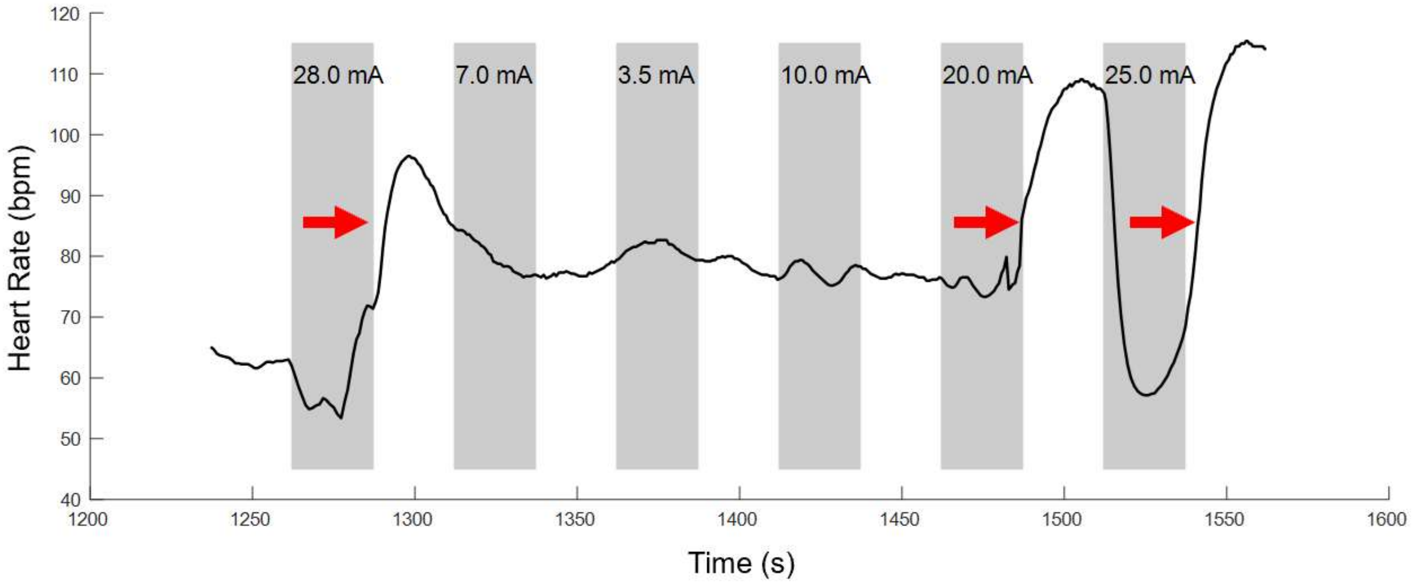


*Figure 30. Participant 6 HR trace from AA00, 50 µs stimulation showing tachycardia occurring after stimulation that evoked bradycardia (28 mA, 25 mA) and after trials that did not evoke bradycardia (20 mA).*
